# Elucidating the causal role of age of menarche, adiposity, lipid fractions, and blood pressure upon cardiovascular disease: A multivariable Mendelian randomization study

**DOI:** 10.1101/2023.06.19.23291629

**Authors:** Yongho Jee, Wes Spiller, Eleanor Sanderson, Kate Tilling, Tom Palmer, Eunhee Ha, YoungJu Kim

## Abstract

This study evaluates the potential role of multiple correlated risk factors upon coronary heart disease (CHD) and ischemic stroke, and the extent to which using GWAS summary data including prevalent cases of stroke, as opposed to incident cases, can influence Mendelian randomization (MR) analyses. Initially, thirteen candidate risk factors were identified through a literature review, including age of menarche, adiposity, blood pressure, lipid fractions, physical activity, type-II diabetes, smoking, sleep duration, alcohol consumption, and kidney function. Using publicly available summary data from genome-wide association studies (GWAS), the total effect of each exposure on CHD, ischemic, and cardioembolic stroke was estimated using univariable summary MR. Multivariable MR (MVMR) analyses were then used to estimate the conditional effects of low-density lipoprotein (LDL), high-density lipoprotein (HDL), triglycerides and systolic blood pressure (SBP) on each outcome. To select the MVMR model a novel forward selection algorithm was applied to include the greatest number of exposures while maintaining sufficient conditional instrument strength for estimation. To examine potential bias from using GWAS summary data derived from prevalent cases of ischemic stroke a GWAS of incident ischemic stroke was conducted using data from the UK Biobank. In univariable MR analyses negative effects of blood pressure were observed across all outcomes, while the effects of remaining exposures differed markedly. HDL was also estimated to have a protective effect on all outcomes except cardioembolic stroke. Univariable and MVMR estimates were directionally consistent, though MVMR estimates were attenuated. Finally, repeating analyses using incident stroke cases yielded results in agreement with prevalent stroke data, suggesting the use of prevalent outcome data did not bias our initial analysis.

## Introduction

Cardiovascular disease (CVD) is a leading contributor to global mortality, responsible for more than 30% of global deaths [1]. As a consequence, identifying modifiable risk factors for CVD related illness has come to represent a global imperative, facilitating the development of strategies for treatment and prevention [1,2]. As CVD encompasses a wide range of illnesses with potentially diverse aetiologies, such a goal is best met through an improved understanding of the causal mechanisms underlying each disease.

With the proliferation of genome-wide association studies (GWAS), summary Mendelian randomization (MR) has emerged as a popular methodological framework for estimating causal effects [3–6]. Summary MR methods utilise genetic variant-phenotype association estimates from publicly available GWAS, producing causal effect estimates which are potentially robust to reverse causation or confounding bias [4,7–9]. While such analyses have often focused on estimating the total effect of a single risk factor on an outcome, the recent development of multivariable summary MR approaches (MVMR) has facilitated evaluation of the conditional effects of multiple exposures simultaneously [10–12]. MVMR approaches are particularly useful when evaluating correlated exposures, providing a more detailed depiction of the causal mechanisms underpinning a given disease.

This study explores the extent to which the causal effects of risk factors for CHD and ischemic stroke are driven by their relationship with additional correlated exposures. Initially, we identify a set of candidate risk factors through literature review as a means of justifying their inclusion in downstream analyses. Causal effects are then estimated for each exposure, with a view to minimising the possibility of selectively reporting results only for risk factors satisfying an arbitrary significance threshold. Following the literature review, we adopted age of menarche, blood pressure, adiposity, lipid fractions, physical activity, type-II diabetes, smoking, alcohol consumption, sleep duration, and kidney function as risk factors for which genetic determinants were plausible.

While it was possible to evaluate the effect of each exposure individually, MVMR estimates could not be reliably obtained when fitting all exposures within a single MVMR model due to insufficient conditional instrument strength. This highlights a common issue when performing MVMR analyses, specifically how to select and justify an appropriate MVMR model when all exposures cannot be used simultaneously. In this paper we propose a novel forward selection algorithm maximising conditional F-statistics for instrument strength to select the greatest feasible number of exposures (see Methods). Assuming that the initial set of risk factors is determined with reference to previous research, such as through literature review, our forward selection approach provides a means through which the greatest number of exposures can be included, while minimising the extent to which exposure selection is arbitrarily determined through researcher biases.

A further issue explored in this paper is potential bias resulting from the use of GWAS summary data from studies using prevalent ischemic stroke cases. Specifically, the MEGASTROKE consortium leverages data from several case-control studies, and as a consequence resulting estimates could be influenced by selection into the study [19]. This could happen in cases where a risk factor is causally related to ischemic stroke severity, such that participants with specific risk factor configurations are more likely to be recruited into the study. For example, if greater levels of alcohol consumption increased ischemic stroke severity, it could be the case individuals with lower levels of alcohol consumption are enriched in the sample of ischemic stroke cases, because increased alcohol consumption increases the extent to which incidents are fatal and only participants surviving an ischemic stroke are included in the study. This can consequently result in reduced alcohol consumption being associated an increased risk of stroke, when it is actually associated with study participation for participants acting as cases.

The extent to which causal effect estimates are driven by the use of GWAS summary data from prevalent ischemic stroke cases is explored by repeating univariable MR and MVMR analyses with GWAS summary data specific to incident ischemic stroke. GWAS summary data is then used in univariable MR and MVMR analyses, comparing resulting estimates to previous analyses using MEGASTROKE GWAS summary data.

## Materials and methods

### Identifying candidate exposures through review

Using the PubMed and Web of Science online databases, the search terms “coronary heart disease”, “myocardial infarction”, “stroke”, “ischemic”, “ischaemic”, ”hemorrhagic”, ”haemorrhagic”, and ”atherosclerosis” were used, searching for instances wherein any of the terms were used (a Boolean ”OR” operator). Author names, article title, year of publication, journal name, and abstracts for each paper were extracted, covering the time period May 2011-May 2021. A three-stage selection process was then conducted, assessing the title, abstract, and main body of each paper sequentially.

Papers were excluded from the review on the basis of five selection criteria:

1. Inaccessible or written in a non-English language.
2. A review or meta-analysis.
3. An animal or cell model study.
4. Unrelated to cardiovascular disease or conducting analyses where controls were defined as having a pre-existing cardiovascular condition, such as hypertension.
5. Of insufficient quality using Joanna Briggs Institute criteria.

A total of 52,931 papers were identified in the initial search, with 2,028 papers remaining after exclusion criteria were applied. With this complete, the subset of 2,028 relevant papers were evaluated with respect to paper quality using the Joanna Briggs Institute (JBI) critical appraisal tools (available at https://jbi.global/critical-appraisal-tools). A summary of the number of papers excluded by category is provided in Fig 1 (left), while a summary of papers identifying statistically significant associations between one or more risk factors and CHD or stroke is presented in Fig 1 (right).

**Fig 1.**
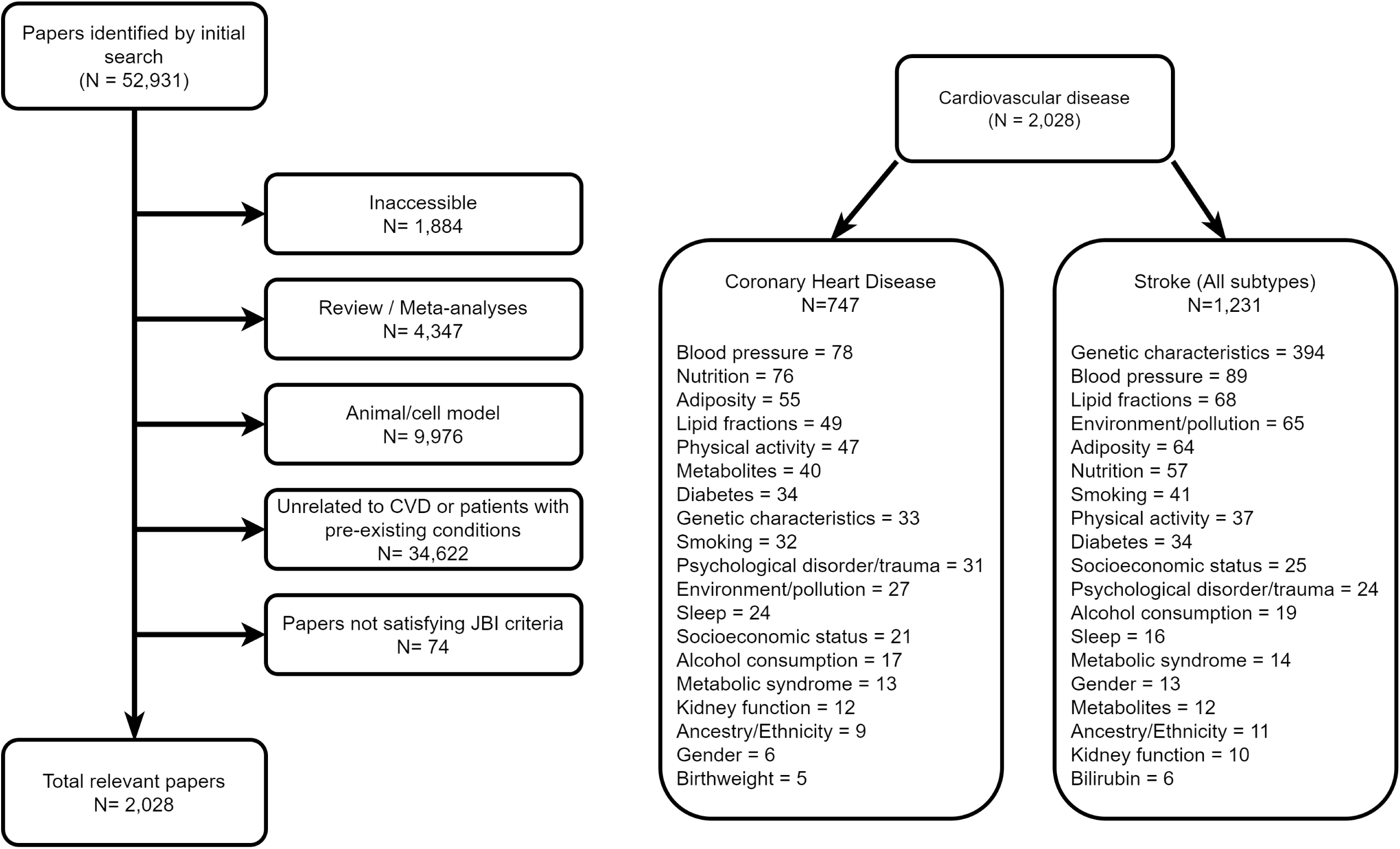
A panel summarizing the papers initially identified through review (left), and number of exposure specific papers for coronary heart disease (CHD) and stroke (right).

The set of remaining papers can further be subcategorised by outcome and risk factor of interest. In Fig 1 (right) a summary of the distribution of papers by outcome of interest (CHD or all subtype stroke) is provided, highlighting sets of frequently examined risk factors. For each paper the presence and direction of significant estimated effects (using a threshold of *p <* 0.05 or corrections for multiple testing where applicable) were extracted. Note that for clarity only exposures for which the number of corresponding papers exceeded five are included in Fig 1. The source documents used for filtering papers are available at https://github.com/WSpiller/CVDMVMR.

Focusing on modifiable risk factors, analyses exploring the role of diastolic blood pressure (DBP) and systolic blood pressure (SBP) on CHD and ischemic stroke were most frequently identified. Systolic and diastolic blood pressure were frequently found to be positively associated with CHD and stroke risk [28–31]. In addition, several studies have suggested that the effect of risk factors such as insulin resistance may vary with changes in blood pressure [32,33]. Adiposity was also identified as a risk factor for both stroke and CHD, with increases in measures such as body mass index (BMI) and waist circumference being estimated to increase disease risk [34–36]. Alcohol consumption, sedentary behaviour, and smoking have also been linked to increased risk of stroke and CHD events [42–47].

A number of studies have highlighted a possible role of serum lipids in CHD and stroke risk, with differing directions of effect depending on the lipid fraction investigated. Broadly, low-density lipoprotein (LDL) and triglycerides are associated with increases in CHD and stroke risk, while high-density lipoprotein (HDL) is frequently estimated to have a protective effect [37–39]. The relationship between metabolites and CHD or stroke has also received substantial attention, though it is often the case that metabolites are considered to represent potential biomarkers for conventional risk factors rather than serving as determinants of CHD and stroke risk [40,41]. In addition, there is a growing body of research focused on the role of sex hormones and development as indicators of future CHD and stroke risk [48,49].

It is interesting to note that competing downstream health conditions such as type-II diabetes and kidney function were identified. Such studies raise questions regarding whether these conditions can adversely impact risk of subsequent CHD or stroke events, potentially in conjunction with traditional CVD risk factors. Finally a set of risk factors were identified for which GWAS summary data is scarce, including environmental, psychological and social exposures. As a consequence, such risk factors are not formally considered in this study, though analyses focusing on these elements of cardiovascular health represent promising avenues for future research.

### Study and variable descriptions

Genetic variant-exposure associations and corresponding standard errors for the majority of candidate risk factors were obtained from the UK Biobank, a cohort study composed of participants aged 40-69 recruited within the United Kingdom [14]. Data for approximately 500,000 individuals was recorded, including anthropometric measures, self-report questionnaires, and biological measures for downstream genotyping [20]. Age of menarche was selected as a proxy variable for hormonal changes in childhood development, recorded in years for female participants. BMI was used as a measure of adiposity, derived using measured of height and weight obtained at baseline. Baseline automated readings for DBP and SBP were used, measured with mmHg units, while LDL, HDL, and triglycerides were recorded in mg/dL from serum measures. In addition, creatinine concentration in urine was used as an indicator of kidney function, measured in mg/dL at baseline. Measures of BMI, DBP, SBP, lipid fractions and creatinine were standardised using a z-score transformation prior to conducting analyses so as for estimates to be interpreted on a standard deviation scale.

The number of days a participant engaged in 10 minutes or more of vigorous physical activity was used as a general measure of physical activity, while sleep duration records the average hours of sleep per day for each participant. Average units of alcohol consumed per week, derived by Clarke et al (2017), and excluding former drinkers was used as a measure of alcohol consumption [21], while smoking behaviour is a categorical variable representing past tobacco smoking. Importantly, as categories represent general smoking behaviour tied to frequency of consumption as opposed to average amount consumed, subsequent estimates are interpreted as representing increases in smoking amount, and are not linked with a specific unit increase. Physical activity, sleep duration, alcohol consumption and smoking behaviour measures were obtained using determined using self-report questionnaire at baseline.

Genetic variant-outcome associations for CHD were sourced from the CARDIOGRAMplusC4D consortium [18]. The CARDIOGRAMplusC4D consortium conducted a large-scale GWAS meta-analysis using information from approximately 180,000 participants across a total of 48 studies. CHD was recorded as a binary variable with approximately 60,800 identified cases. Incident CHD cases were defined using International Classification of Diseases 9th Revision (ICD-9) codes 410-414, and ICD-10 codes I20-I25. A sample of mixed ancestry was used, though participants were predominantly of European ancestry (77%). For ischemic stroke summary data were obtained from the MEGASTROKE consortium, including large-artery atherosclerosis (4,373 cases), cardioembolic (7,193 cases), and small vessel (5,386 cases) subtypes as defined by the Trial of Org 10172 in Acute Stroke Treatment (TOAST) criteria [13]. MEGASTROKE participants were primarily of European ancestry.

For analyses restricted to incident stroke, cases were identified using HES data within the UK Biobank, generating a binary variable indicating one or more stroke events occurring after baseline measures were recorded. Individuals having previously experienced a stroke event were excluded, and stroke cases were identified using ICD-10 codes I61-6363. Resulting estimates represent effect of each exposure on general ischemic stroke as opposed to specific ischemic stroke subtypes.

### Univariable and Multivariable Mendelian randomization

MR uses genetic variants, often single nucleotide polymorphisms (SNPs), as instrumental variables to estimate causal effects which are not biased by unmeasured confounding. This is possible in cases where a candidate genetic variant is strongly associated with an exposure of interest (IV1), there are no confounders of the genetic variant and the outcome (IV2), and independent of the outcome when conditioning on the exposure (IV3) [22]. These conditions are illustrated in Fig 2 for a genetic variant *G*, exposure *X*_1_, outcome *Y*, and set of unmeasured confounders *U*.

**Fig 2.**
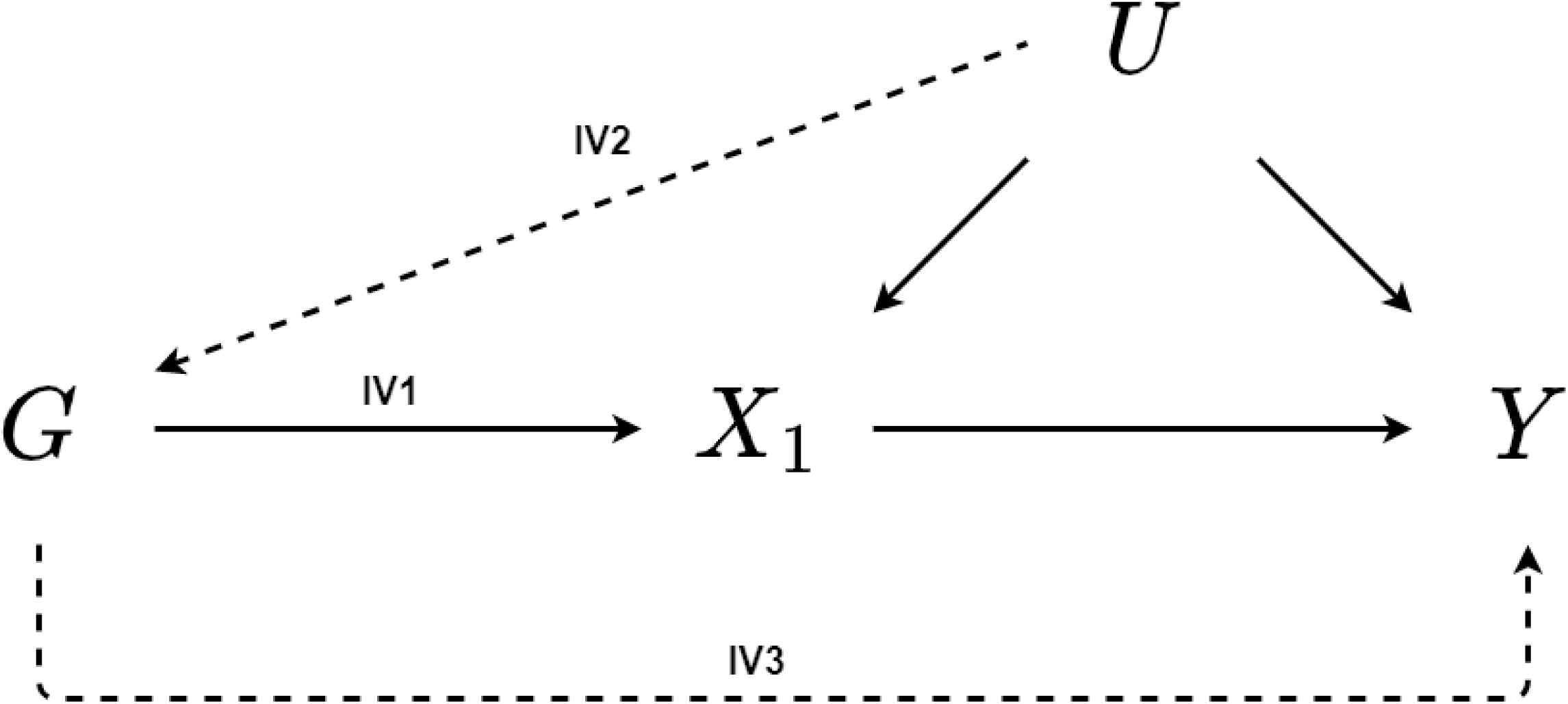
A directed acyclic graph depicting the assumed relationships for unbiased inference using MR. The arrow between the genetic instrument *G* and exposure *X*_1_ represents a strong association satisfying assumption IV1. Dashed lines represent associations required to be zero in order for assumptions IV2-3 to be satisfied.

For an individual genetic instrument, a Wald ratio estimate can be obtained by dividing the genetic variant-outcome association by the genetic variant-exposure association [4,23]. Where multiple SNPs are available, ratio estimates are often combined through fixed-effects meta-analysis [4,7]. The inverse variance weighted (IVW) estimate represents an average of multiple ratio estimates, weighted by the inverse of the variance of the genetic variant-outcome association.

Violation of the IV assumptions can introduce bias into causal effect estimates. Assumption IV1 can be tested through an F-statistic, however it is not possible to formally test assumptions IV2 and IV3. However, provided associations violating IV3 are not identically distributed across the range of instruments used, heterogeneity in individual effect estimates can serve as an indicator of IV3 violation [24]. Heterogeneity can be estimated using Cochran’s Q-statistic, such that genetic instruments contributing substantially to global heterogeneity are typically identified as invalid [25].

In cases where one or more genetic variants are invalid several methods for obtaining MR estimates that are robust to that violation are available. MR-Egger regression allows for directional pleiotropy by extending the IVW regression model to include an intercept, where a non-zero intercept is interpreted as an average of pleiotropic effects across the set of genetic variants [7]. This hinges upon an additional assumption that the size of the pleiotropic effect for each SNP is independent of the strength of that SNP as an instrument. The weighted median approach selects the median per SNP ratio estimate with respect to weighting, providing an unbiased causal effect estimate where the weighted proportion of valid genetic variants exceeds 50% [8]. The weighted modal estimator assumes that the modal ratio estimate *β_j_* represents estimates obtained from valid genetic variants. This is referred to as the Zero Modal Pleiotropy Assumption (ZEMPA) [9]. This will give an unbiased effect estimate when the largest group of SNPs is not pleiotropic.

MVMR extends the univariable MR framework to contain multiple exposures simultaneously [11,12], adjusting for each exposure to provide estimates of direct effect. The assumptions of MVMR build upon the existing MR assumptions, with MVMR relying upon genetic variants:

1. Strongly associated with each exposure when conditioning on remaining exposures (MVMR1).
2. No confounding of any of the genetic variants and the outcome (MVMR2)
3. Independent of the outcome when conditioning on all included exposures (MVMR3)

Assumption MVMR1 is assessed by calculating the conditional F-statistic for each exposure, typically using a threshold value of 10 to indicate sufficient instrument strength for conducting analyses [12,26]. Violations of MVMR3, resulting from continued invalidity after controlling for additional exposures can be evaluated by calculating an adjusted Q-statistic, as described in Sanderson et al [12]. Crucially, the inclusion of additional exposures has the potential to address the limitations of the univariable summary MR approaches previously discussed. Specifically, comparing univariable and conditional F-statistics can highlight the extent to which genetic variants are predictive for multiple exposures, resulting in potential invalidity.

### MVMR model selection

To select an appropriate MVMR model with respect to instrument strength, a novel forward selection algorithm is presented with the aim of including the highest number of exposures for which conditional F-statistics exceed a threshold value of 10. Initially, all possible pairs of exposures are evaluated with respect to their conditional F-statistics. The mean F-statistic for each pair is then calculated, weighted by the inverse of the absolute value of the difference between F-statistics within a given pair. The rationale behind weighting the mean F-statistic for each pair stems from instances where two exposures have a high mean F-statistic, driven primarily by one of the exposures. For example, consider two pairs of exposures *A* and *B* where each pair has a mean conditional F-statistic of 50. Further, assume the conditional F-statistics for pair *A* and pair *B* are (*F*_1_ = 10*,F*_2_ = 90) and (*F*_1_ = 50*,F*_2_ = 50) respectively. In this case, the similarity in mean conditional F-statistic could mistakenly imply that the conditional F-statistics for both pairs are similar.

In the above example the decision to prefer pair *B* over pair *A* is arbitrary, given that the conditional F-statistics for all exposures exceeds the commonly used threshold of 10. The distinction is important, however, when considering the inclusion of additional exposures within the MVMR model. A conditional F-statistic reflects the strength of association between a set of genetic instruments and a given exposure, conditioning on additional exposures included within the MVMR model. Consequently, if an exposure is added which serves as a mediator between an existing exposure and one or more of its corresponding genetic instruments, it would be follow that the conditional F-statistic for the existing exposure would decrease. Essentially, a proportion of the instrument strength for the existing exposure can be attributed to the relationship between the two exposures, and is removed when conditioning on the additional exposure. Returning to hypothetical exposure pairs *A* and *B*, pair *B* can more easily accommodate reductions in F-statistic while satisfying the threshold of 10 in contrast with pair *A*.

Before continuing it is important to highlight two key considerations when using conditional F-statistics for MVMR model selection. First, the inclusion of an additional exposure does not necessarily imply conditional F-statistics for the existing exposures will decrease. Conditional F-statistics are most likely to decrease when the additional exposure lies on the causal pathway from the genetic variants to an existing exposure. Second, when conditional F-statistics are found to decrease, and the candidate additional exposure is omitted from the model, resulting direct effect estimates for the remaining exposures implicitly include the effect of the omitted exposure on the outcome. If the effect of the omitted exposure is entirely mediated through the existing exposures, or the effect of the existing exposures is mediated entirely through the omitted exposure, then omitting the additional exposure will not introduce bias into MR estimates. However, where the omitted exposure exerts a direct effect on the outcome the resulting MR estimates will be biased in the direction of this association. These two scenarios describe vertical and horizontal pleiotropic relationships respectively.

### Statistical analysis

For MR analyses sets of independent genetic variants were initially for each exposure using the MR-Base online GWAS database [17,27]. A p-value threshold of *p <* 5 × 10^−8^ was used to identify genetic variants robustly associated with the phenotype of interest, and linkage-disequilibrium (LD) pruning was implemented using a threshold of *r*^2^ *<* 0.01 and a clumping window of 10,000 kb to ensure only independent genetic variants were selected. For each outcome univariable MR analyses were performed, specifically using IVW, MR-Egger, weighted median, and weighted modal approaches.

To select an appropriate MVMR model we initially calculated the weighted mean conditional F-statistic for each exposure pair. With this complete, we selected the pair with the highest weighted mean conditional F-statistic and constructed a set of models by including one of the remaining exposures individually. This process was iterated until remaining exposures either failed to exceed the given threshold, or the inclusion of an additional exposure would lower the conditional F-statistic for any included exposure below the instrument strength threshold. Ideally it would be desirable to include all potentially relevant exposures within a single MVMR model, yet it was not possible to do so while maintaining sufficient conditional instrument strength. The final model used in MVMR analyses therefore reflects a trade-off between including modifiable risk factors of interest and producing causal estimates of sufficient precision so as to be informative. GWAS association estimates were obtained from the MR-base GWAS database, and analyses were performed using the TwoSampleMR, RadialMR, and MVMR R packages [12,25,27]. Analyses were conducted in R (version 4.0.3), and code for the analyses is available at https://github.com/WSpiller/CVDMVMR.

To evaluate potential bias from using data derived from studies using prevalent stroke cases, we conducted a GWAS using genetic and HES data from UK Biobank using information on unrelated participants of European ancestry. Outcome data was sourced from the HES database, extracting date of hospital admission and corresponding diagnoses categorised using ICD-10 codes. Initially participants with a history of ischemic stroke prior to recruitment were removed, corresponding to ICD-10 codes I61-I63. Individuals experiencing a stroke event after followup were then categorised as cases (*N* = 9,527). Finally, a random subset of individuals was selected as controls, maintaining a case to control ratio of 1:10 and a total sample size *N* = 95,270. Effects were obtained using BOLT-LMM adjusting for participant age and sex in addition to array batch effects. Resulting estimates and standard errors were transformed so as to be interpreted as a log-odds ratio, and descriptive statistics for the GWAS sample are provided in supplementary Table S1.

## Results

In univariable MR analyses thirteen sets of genetic instruments were obtained, corresponding to each of the exposures previously described. A total of 2038 SNPs were used across all exposures, with mean F-statistics across all analyses ranging from 36 to 177. Summary data for each univariable analysis available at https://github.com/WSpiller/CVDMVMR, in addition to code for downloading and replicating each analysis. Fig 3 shows a forest plot displaying the IVW estimate and 95% confidence interval for each exposure, grouped by outcome. Effect estimates have been exponentiated so as to be interpreted on an odds ratio scale. To account for multiple testing a p-value of *p <* 0.05*/*13 was used as a threshold for statistical significance.

**Fig 3.**
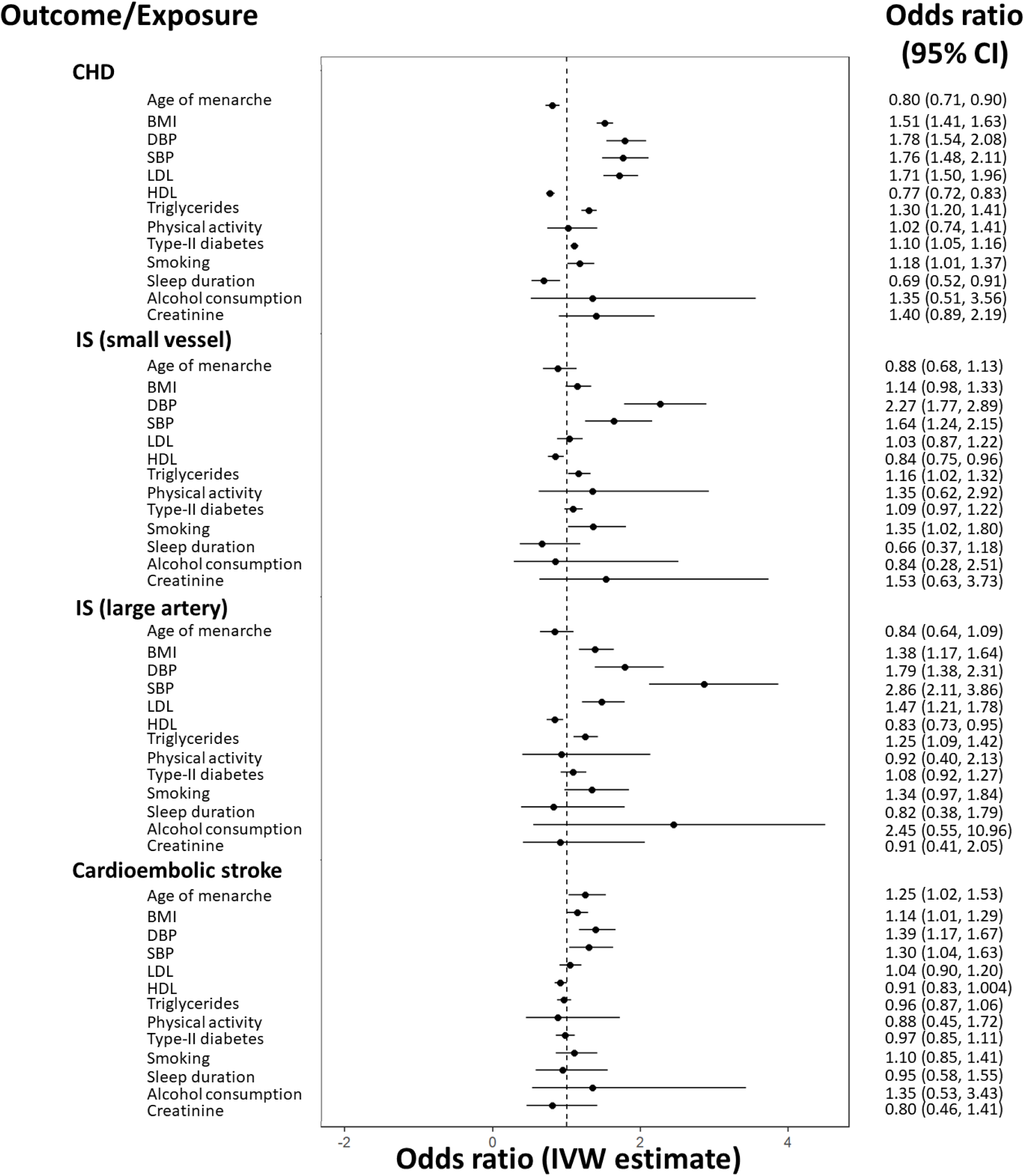
A forest plot showing IVW univariable MR estimates. Estimates are grouped by outcome. Effect estimates on the odds ratio scale, with accompanying 95% confidence intervals are illustrated, with corresponding values shown to the right of the plot.

For CHD seven exposures showed evidence of a positive effect, including BMI, DBP, SBP, LDL, triglycerides, type-II diabetes and smoking. BMI, blood pressure and LDL exert effects of similar magnitude, representing change in CHD risk per standard deviation increase. Genetically predicted type-II diabetes status was found to increase the risk of CHD by 10% (OR=1.10, 95% CI=1.05, 1.16), while increased frequency of tobacco consumption was positively associated with CHD risk (OR=1.18, 95% CI=1.01, 1.37). Later onset of menarche (OR=0.80, 95% CI=0.71, 0.90), HDL (OR=0.77, 95% CI=0.72, 0.83) and increased sleep duration (OR=0.69, 95% CI=0.52, 0.91) were found to be protective with respect to CHD. Estimates for increased concentration of creatinine, representing reduced kidney function, were suggestive of a positive effect on CHD risk for which the analyses was under-powered (OR=1.40, 95% CI=0.89, 2.19). Similarly, the wide confidence interval for alcohol consumption reflects a lack of statistical power to detect a causal effect on CHD. Physical activity showed no evidence of a causal effect on CHD (OR=1.02, 95% CI=0.74, 1.41).

Considering small vessel ischemic stroke DBP (OR=2.27, 95% CI=1.77, 2.89) and SBP (OR=1.64, 95% CI=1.24, 2.15) appear to have the most substantial effects. BMI, triglycerides, type-II diabetes and smoking behaviour showed evidence of positive associations with small vessel ischemic stroke risk, while HDL appeared to have a protective effect (OR=0.84, 95% CI=0.75, 0.96). LDL showed no evidence of a directional effect on small vessel ischemic stroke (OR=1.03, 95% CI=0.87, 1.22). Sleep duration showed limited evidence of a protective effect for which the analyses were under-powered (OR=0.66, 95% CI=0.37, 1.18), while power constraints limited the extent to which physical activity, alcohol consumption or creatinine could be considered.

Comparing small vessel and large artery ischemic stroke subtypes, blood pressure again exerted the greatest effects on large artery ischemic stroke risk. The positive effects of BMI (OR=1.38, 95% CI=1.17, 1.64) and triglycerides (OR=1.25, 95% CI=1.09, 1.42) on large artery ischemic stroke were greater in magnitude, while smoking behaviour and HDL yielded effects in agreement with small vessel ischemic stroke. Notably, LDL is estimated to have a substantial positive effect on large vessel ischemic stroke (OR=1.47, 95% CI=1.21, 1.78), while the effect of type-II diabetes was attenuated (OR=1.08, 95% CI=0.92, 1.27). Remaining exposures had insufficient precision to identify causal effects.

For cardioembolic stroke DBP was the only exposure with an observed association after correcting for multiple testing (OR=1.39, 95% CI=1.17, 1.67). Estimates for age of menarche show evidence of a positive association with cardioembolic stroke (OR=1.25, 95% CI=1.02, 1.53) in contrast with the protective effect observed for CHD. Estimates for BMI and SBP seem to indicate potential roles as risk factors, in addition to a protective effect of HDL (OR=0.91, 95% CI=0.83, 1.004). LDL, physical activity, sleep duration, tobacco consumption and creatinine showed no evidence of a directional causal effect, while analyses of alcohol consumption lacked statistical power.

Overall, the results depicted in Fig 3 highlight key differences in potentially relevant risk factors across the range of outcomes considered. Age of menarche seems related to CHD and cardioembolic stroke, though the direction of causal effect differs. BMI and blood pressure are consistently highlighted as risk factors exerting a positive effect on all outcomes, while LDL and triglycerides appear to only influence CHD and large artery ischemic stroke. HDL is estimated to have a protective effect with respect to the range of outcomes, while a positive effect of type-II diabetes was only observed for CHD. Smoking was a risk factor for all outcomes except for cardioembolic stroke, while estimates indicative of a causal role of sleep duration and kidney function were only found for CHD. Unfortunately, analyses of alcohol consumption and physical activity were not sufficiently precise to identify potential directional effects.

The sets of univariable estimates presented were obtained using the IVW method, and as a consequence can be biased where horizontal pleiotropic pathways are present (violations of IV3). Using Cochran’s Q-statistic as an indicator of potential pleiotropic bias, substantial heterogeneity was only present for age of menarche when CHD served as an outcome (Q=205.7, *p <* 0.001). Heterogeneity appeared to be present in all BMI and blood pressure analyses, and for lipid fractions all analyses showed evidence of heterogeneity with the exception of HDL (Q=313.49, *p* = 0.561) and triglycerides (Q=269.43, *p* = 0.600) in relation to cardioembolic stroke. Analyses focusing on physical activity and type-II diabetes did not show evidence of substantial heterogeneity, while smoking (Q=156.8, *p <* 0.001) and alcohol consumption (Q=14.9, *p <* 0.001) only exhibited heterogeneity when considering CHD. Creatinine had evidence of heterogeneity in analyses of CHD (Q=47.95, *p <* 0.001) and small vessel ischemic stroke (Q=35.59, *p* = 0.027). Heterogeneity statistics for each analysis are provided in supplementary Table S2.

In cases where heterogeneity was observed, invalid instruments were identified and removed based on their contribution to global heterogeneity. This was achieved by evaluating the Q-statistic for each instrument individually, using a p-value threshold with a multiple testing correction equal to 0.05 divided by the total number of instruments in the IVW analysis. IVW analyses performed using sets of instruments following outlier removal, in addition to applying sensitivity analyses, produced results in agreement with the IVW estimates presented in Fig 3 (see Supplementary Material Table S3).

In MVMR analyses it was not possible to fit all 13 exposures simultaneously while maintaining sufficient conditional instrument strength to estimate causal effects. For example, fitting a six exposure MVMR model yielded conditional F-statistics for age of menarche (*F* ≈ 11), BMI (*F* ≈ 6), SBP (*F* ≈ 9), LDL (*F* ≈ 4), HDL (*F* ≈ 2), or triglycerides (*F* ≈ 2). As previously discussed, the attenuation of conditional F-statistics when fitting additional exposures could be due to either a low proportion of the total set of SNPs being associated with a particular exposure or because the set of genetic variants is predictive of multiple exposures (see Methods) [12].

As a consequence, effect estimation using MVMR required a subset of exposures to be used. Using the forward selection procedure previously described (see Methods), all potential pairs of exposures were evaluated, with HDL and triglycerides providing the highest weighted sum of conditional F-statistics. After evaluating additional exposures sequentially LDL and SBP were also included in a four exposure MVMR model. Fig 4 is a heat map presenting the conditional F-statistic for each exposure (labelled exposure 1), and illustrating the magnitude of the weighted mean conditional F-statistic as a gradient.

**Fig 4.**
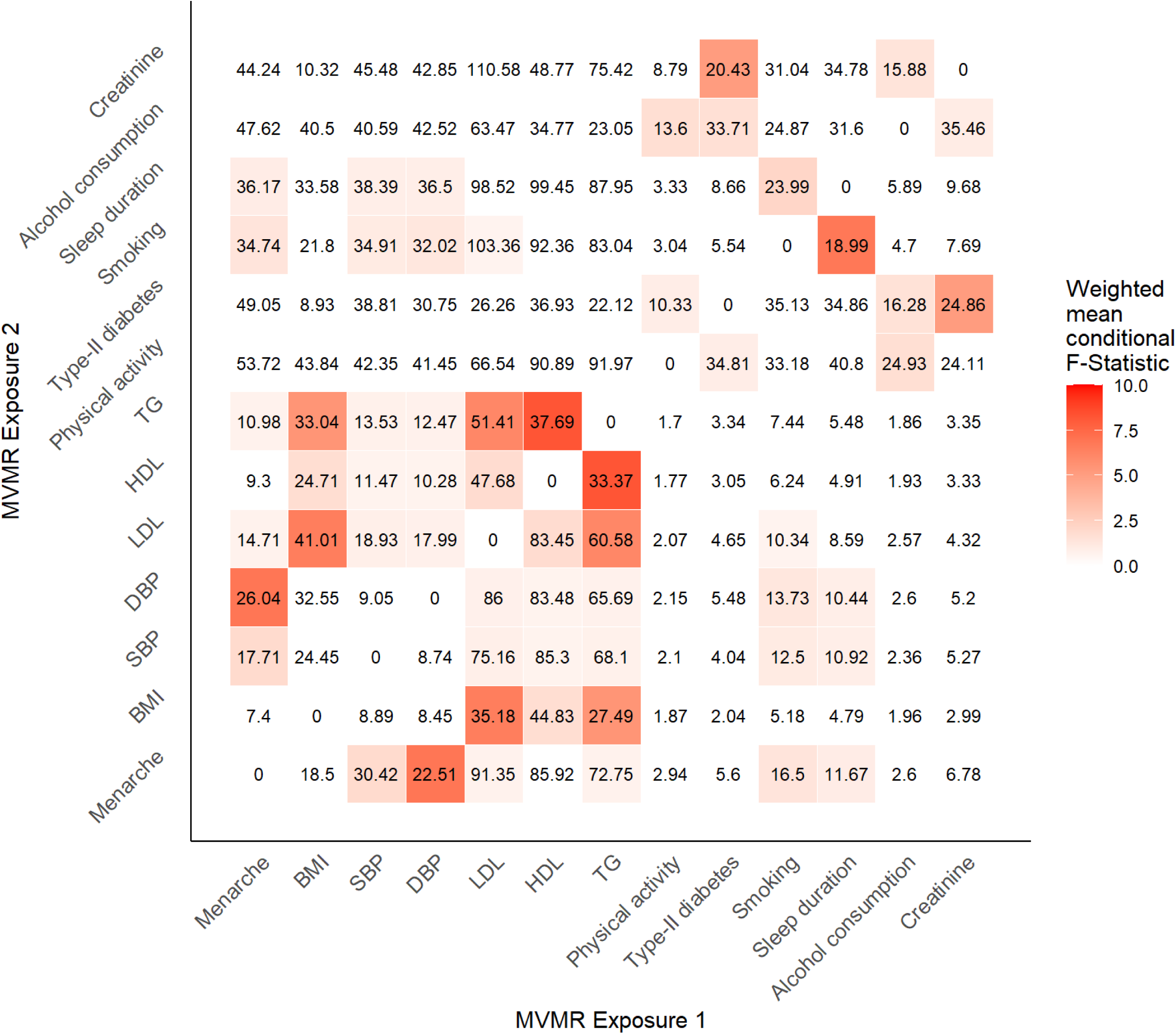
A heatmap showing the conditional F-statistic for each exposure when included in a two variable MVMR model. The conditional F-statistic corresponds to the exposure labelled on the x-axis when fit in a two variable MVMR model including the corresponding exposure on the y-axis. A colour gradient is used to indicate the magnitude of the weighted mean conditional F-statistic.

When including LDL, HDL, triglycerides, and SBP in the MVMR model the number of instruments differed across the range of outcomes. This reflects differences in the availability of outcome-specific GWAS summary data for specific SNPs across selected GWAS. Specifically, a total of 1261 instruments were used to estimate effects on CHD, 1296 for small vessel ischemic stroke, 1300 for large artery ischemic stroke and 1291 for cardioembolic stroke. Fig 5 shows the estimates obtained using all exposures simultaneously, with superimposed univariable IVW estimates for ease of comparison.

**Fig 5.**
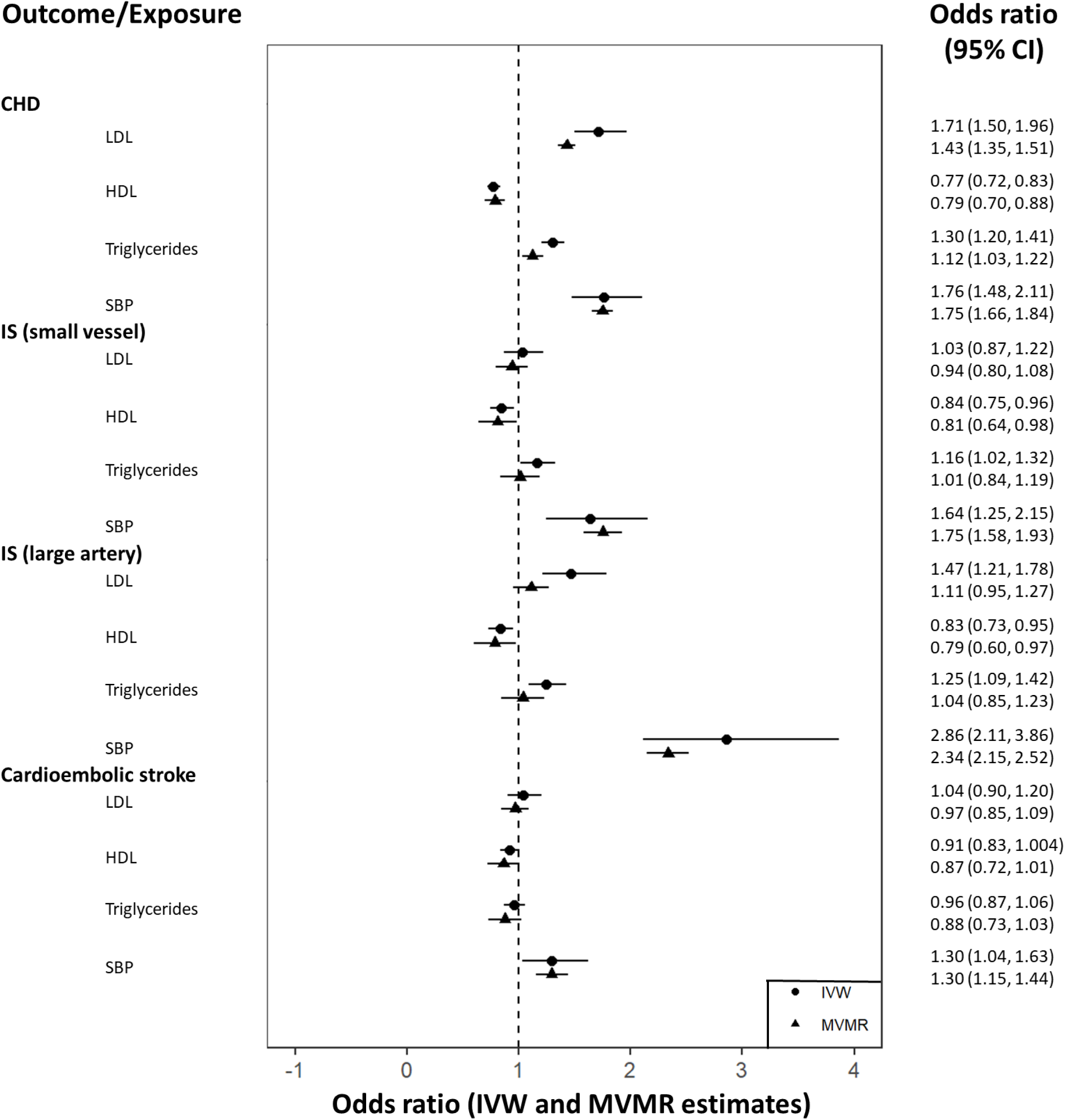
A forest plot showing MVMR MR estimates. Estimates are grouped by outcome, representing the direct effect of each exposure including all exposures within each MVMR model. Effect estimates on the odds ratio scale, with accompanying 95% confidence intervals are illustrated, with corresponding values shown to the right of the plot. Univariable MR estimates for each exposure are included for comparison.

When adjusting for multiple exposures, LDL (OR=1.71, 95% CI=1.50, 1.96), triglycerides (OR=1.12, 95% CI=1.03, 1.22), and SBP (OR=1.75, 95% CI=1.66, 1.84) were positively associated with CHD. The effects of LDL and triglycerides were attenuated but directionally consistent with univariable MR estimates. HDL and SBP were found to be in agreement with univariable MR estimates, with a substantial improvement in precision for SBP. The observed agreement in effect estimates for SBP suggests previous univariable estimates are unlikely to be driven by genetic effects shared between exposures included in the MVMR model, though the attenuation visible for LDL and triglycerides suggests that there may be some overlap in genetic effects in univariable analyses. This could be interpreted as an instance where effects are greater due to horizontal pleiotropic effects.

For small vessel ischemic stroke a protective effect of HDL similar in magnitude to the corresponding univariable MR estimate was observed (OR=0.81, 95% CI=0.64, 0.98). SBP was again estimated to have a positive effect on small vessel ischemic stroke (OR=1.75, 95% CI=1.58, 1.93), while the effect of LDL remained negligible (OR=0.94, 95% CI=0.80, 1.08). Comparing the univariable and MVMR estimated effects of triglycerides, the positive association with small vessel ischemic stroke is not observed when adjusting for LDL, HDL and SBP (OR=1.01, 95% CI=0.84, 1.19). This would suggest that the positive association observed in the univariable analyses may be driven by associations with the additional exposures included in the MVMR model.

Considering large artery ischemic stroke estimates for HDL (OR=0.79, 95% CI=0.60, 0.97) and SBP (OR=2.34, 95% CI=2.15, 2.52) remain directionally consistent with the corresponding univariable MR estimates. SBP continues to have a more substantial effect on large artery ischemic stroke in comparison with small vessel ischemic stroke, and a substantial increase in precision is observed for both stroke subtypes. When adjusting for additional exposures in the MVMR model, the positive associations of triglycerides and LDL are greatly attenuated to include the null with respect to large artery ischemic stroke (OR=1.11, 95% CI=0.95, 1.27).

Finally, estimates in relation to cardioembolic stroke were largely in agreement with previous univariable MR estimates. A evidence of a protective effect of HDL is observed (OR=0.87, 95% CI=0.72, 1.01), while the positive effect of SBP on cardioembolic stroke risk remains similar in magnitude to the univariable MR estimate (OR=1.30, 95% CI=1.15, 1.44).

### Analyses using incident ischemic stroke from UK Biobank

After conducting a GWAS of incident stroke using individual-level data from the UK Biobank, univariable MR and MVMR analyses were performed using GWAS summary data specific to incident stroke. Fig 6 shows resulting estimates, including MVMR estimates using LDL, HDL, triglycerides and SBP. Comparisons with prevalent ischemic stroke are made specifically with reference to small vessel and large artery ischemic stroke subtypes. A subset of GWAS summary data for incident stroke, including all variants selected as instruments in univariable MR and MVMR analyses is available online at https://github.com/WSpiller/CVDMVMR.

**Fig 6.**
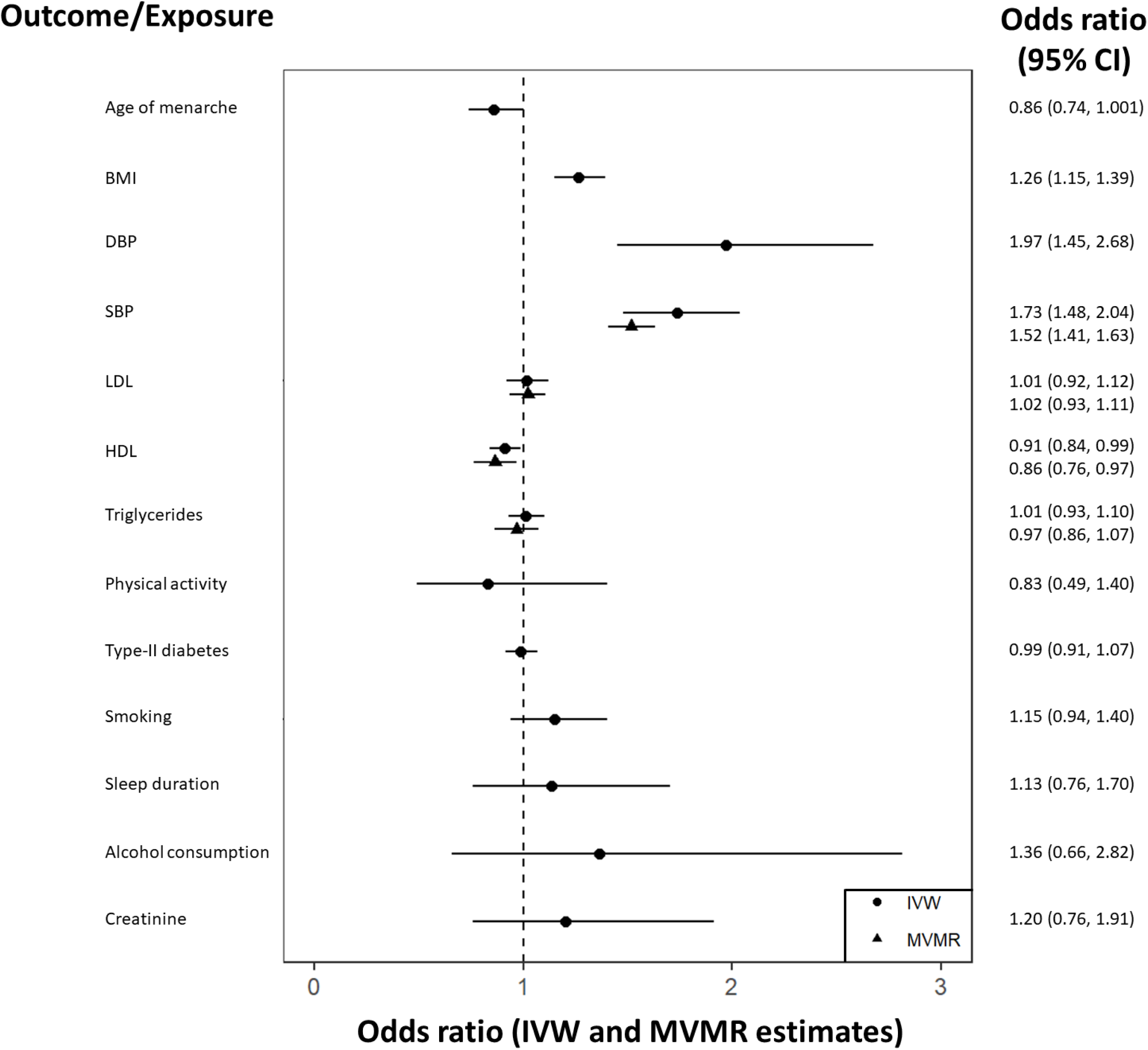
A forest plot showing IVW univariable and MVMR MR estimates for incident stroke using data from the UK Biobank. Effect estimates on the odds ratio scale, with accompanying 95% confidence intervals are illustrated, with corresponding values shown to the right of the plot.

Evaluating univariable MR and MVMR estimates in relation to incident ischemic stroke LDL, HDL, triglycerides, and SBP remain directionally consistent. As was the case for prevalent stroke, the precision of the estimated effect of SBP is substantially improved (OR=1.52, 95% CI=1.41, 1.63), while LDL and triglycerides exhibit negligible effects. Further, the protective effect of HDL remains present after adjustment for additional exposures (OR=0.86, 95% CI=0.76, 0.97).

When comparing estimates using prevalent and incident stroke summary data an initial observation is the more pronounced protective effect of age of menarche on ischemic stroke risk (OR=0.86, 95% CI=0.74, 1.001). BMI, DBP, and SBP are again identified as risk factors for incident ischemic stroke, with blood pressure again exerting the most substantial effect. The protective effect of HDL (OR=0.86, 95% CI=0.76, 0.97) on incident ischemic stroke remains consistent analyses using prevalent stroke data, as well as the positive effect of smoking behaviour (OR=1.15, 95% CI=0.94, 1.40).

A notable difference between the incident stroke and prevalent stroke analyses is the estimated effects of LDL and triglycerides. Using prevalent stroke data, a positive effect of triglycerides on small vessel and large artery stroke was observed, while a positive effect of LDL was only observed for large artery ischemic stroke. Assuming the univariable MR estimate for LDL is specific to large artery stroke, one explanation could be that the general ischemic stroke measure used for incident cases is enriched for small vessel ischemic stroke. This would have the effect of producing an average causal effect weighted by the ratio of ischemic stroke subtypes included in the general measure of ischemic stroke. This could also explain the discrepancy with triglycerides, where the effect is specific to stroke subtypes within the general measure of incident ischemic stroke. However, when considering MVMR estimates the effects of LDL and triglycerides appear negligible. If the observed associations from univariable MR analyses are driven by associations with exposures included in the MVMR model, then estimates subject to ischemic stroke subtype-specific pleiotropic bias are likely of limited utility in practice. The broad agreement in findings using incident and prevalent ischemic stroke data suggests that this feature of the MR analyses is not driving observed causal effects.

## Discussion

This study has explored the potential role of thirteen candidate risk factors on CHD and subtypes of ischemic stroke. Evaluating the role of age of menarche using univariable summary MR, there was evidence of a negative association with CHD, such that menarche occurring at an early age was protective against CHD. This is interesting given that age of menarche is positively associated with relative body size and rate of aging, potential risk factors for CHD [50]. As highlighted by review, such an association could result from fluctuations in hormone levels, in a similar fashion to links between CHD and menopause. One potential explanation for this could be reduced exposure to biological changes related to childhood development, for example exposure to ovarian oestrogen, over the life course. However, when using an MVMR model instrument strength was insufficient to evaluate causal effects, particularly for age of menarche where the sample size for the GWAS was smaller due to analyses only including female participants. This implies that included risk factors are linked to age of menarche through shared genetic pathways.

Similarly, BMI was identified in univariable analyses as a risk factor for CHD and large artery ischemic stroke, yet was not included in the MVMR model due to insufficient conditional instrument strength. As such, if age of menarche or BMI exert direct effects upon CHD independent of lipid fractions or SBP MVMR estimates could exhibit pleiotropic bias through horizontal pleiotropy. It is, however, also possible that age of menarche and BMI lay on the causal pathway between lipid fractions or SBP and the outcomes considered, though this does not seem biologically plausible as it seems unlikely SBP would be causally upstream of BMI. Such a scenario, termed vertical pleiotropy, would not necessarily result in biased effect estimates.

Blood pressure was consistently found to have a positive causal effect on all outcomes across univariable and MVMR analyses, echoing findings from previous analyses [51]. MVMR analyses exhibited a notable increase in estimate precision, which can be explained a reduction in univariable MR heterogeneity when adjusting for lipid fractions. Positive effect estimates of blood pressure on all forms of ischemic stroke highlight an area for effective intervention, demonstrated through the effectiveness of blood pressure lowering medication such as ACE inhibitors with respect to stroke prevention [52].

A protective effect of HDL was frequently observed with respect to ischemic stroke, suggesting it may serve as a potential target for ischemic stroke prevention. It is, however, important to note that such effects have not been observed in randomized control trials evaluating the effectiveness of HDL-raising drugs. For example, a systematic review conducted by Kaur et al (2014) highlights an apparent a lack of evidence of a protective effect of HDL with respect to all-cause mortality and secondary outcomes including ischemic stroke [53]. This study also considered the use of prevalent stroke cases in MEGASTROKE as a potential explanation for the observed protective effect of HDL, motivating the use of incident stroke data from the UK Biobank. The protective effect remained consistent when restricting analyses to data using incident stroke cases, suggesting that if HDL is not causally relevant other mechanisms may be responsible such as relationships between HDL levels and statin use [54].

Sleep duration was found to have a protective effect on CHD, while the impact of sleep on ischemic stroke subtypes was negligible. It should be noted, however, that the effect of sleep on small vessel ischemic stroke was indicative of a potential protective effect for which statistical power was insufficient. Similarly, increased tobacco consumption had marginal effects on CHD, small vessel ischemic stroke and large artery ischemic stroke. Rather than relying on p-value thresholds alone to highlight causal effects it can in many cases be prudent to consider such estimates with respect to their relative precision. Where indicated, in cases where confidence intervals corresponding to an estimate are noticeably wide, a more conservative interpretation is that there may be an effect which could be identified with improvements in sample size. One example where such an effect seems plausible is the univariable MR estimate for creatinine on CHD, which appears suggestive of a positive effect. It is with these limitations in mind that effects which border classic statistical significance thresholds have been highlighted, rather than relying on p-value thresholds alone [55].

Using summary MR approaches affords this study a number of strengths. Initially, the large study samples of UK Biobank, CARDIOGRAMplusC4D, and MEGASTROKE afford substantial statistical strength to identify causal effects. It is important to note, however, that large-scale studies with GWAS summary data related to the exposures and outcomes of interest are still relatively limited. As a consequence, it was not possible to identify genetic instruments in an external independent sample, and it is possible that instrument-exposure associations suffer from Winner’s curse. This describes a situation wherein observed associations occur due to specific features of a sample, and are subsequently unlikely to be replicated in alternative GWAS. With increases in study sample sizes, future work could leverage improvements in instrument strength to include all risk factors, allowing for the adjustment of potential pleiotropic pathways.

An additional strength of this study is the use of a two-sample study design when conducting summary MR analyses. Such a study design is necessary for the application of sensitivity analyses such as MR-Egger regression, and in the univariable MR setting ensures results more conservative when weak instruments are utilised. Though findings were consistent when applying sensitivity analyses in the univariable MR setting, substantial heterogeneity served as an indicator for potential violations of the MR assumptions, warranting further examination using MVMR.

A novel aspect of this work is the use of a novel model selection algorithm for MVMR, implemented by maximising weighted conditional F-statistics across candidate exposures. While an initial set of candidate exposures is best identified with reference to previous research, use of such an approach can prove helpful when selecting a subset of exposures from a range of equally viable risk factors. Code for implementing the method and replicating study findings is available at https://github.com/WSpiller/CVDMVMR.

One limitation of this study relates to the source of GWAS summary data for univariable analyses evaluating age of menarche as an exposure. Due to a lack of available data, outcome GWAS were not sex specific, and as a consequence it is possible that the inclusion of male participants attenuates variant-outcome associations towards the null. Essentially, the contribution of a genetic instrument to the outcome of interest through age of menarche will necessarily not be present in male participants. Use of GWAS summary data including male and female participants has relied on adjustment for participant sex in the GWAS to account for differences in the effect of genetic variants on their respective traits. For example, in the GWAS of BMI, adjustment for participant sex essentially removes the contribution of participant sex to the association estimate. With increases in sample size and available GWAS summary data an evaluation of the impact of using sex-specific data represents a promising area for future research.

A final limitation of this study is the use of a general ischemic stroke measure when conducting analyses of incident stroke. This was motivated by a lack of ischemic stroke subtype-specific cases with which to conduct GWAS and implement downstream summary MR approaches. With the continued proliferation of large-scale population based cohorts with genetic data and hospital record linkage, it is hoped that these analyses can be revisited with a view to identifying unique aspects of the causal mechanisms underpinning different subtypes of ischemic stroke.

## Supporting information

Additional information, including code for conducting analyses and review materials, is available at https://github.com/WSpiller/CVDMVMR.

## Data Availability

The paper predominantly uses GWAS summary data which is publicly accessible from https://www.mrbase.org/, and extracted using code provided. In addition the paper uses GWAS summary data obtained using incident stroke data from the UK Biobank. Summary data for this GWAS is available online at https://github.com/WSpiller/CVDMVMR, including code for replicating analyses. Access to individual data needed to conduct a GWAS of incident ischemic stroke can be obtained by submitting an application to https://www.ukbiobank.ac.uk/enable-your-research/apply-for-access.

## Acknowledgments

The authors would like to thank participants of the UK Biobank from which individual-level data was sourced. Analyses were performed under application number 81499.

